# Assessing how women access healthcare to inform cervical cancer and HIV screening in rural Uganda

**DOI:** 10.1101/2024.10.22.24315934

**Authors:** Mia Sheehan, Hallie Dau, Maryam AboMoslim, Priscilla Naguti, Nelly Mwandacha, Amy Booth, Candice Ruck, Laurie Smith, Jackson Orem, Gina Ogilvie, Carolyn Nakisige

**Author notes:** **Corresponding author:** Gina Ogilvie, MD MSc FCFP DrPH, BC Women’s Hospital and Health Centre, Box 42, Room H203G - 4500 Oak Street, Vancouver, BC, V6H 3N1. **Author contributions:** MS (formal analysis, writing—original draft, writing—review and editing); HD (formal analysis, writing—original draft, writing—review and editing); MA (writing—review and editing); PN (project administration, data collection, writing—review and editing); NM (formal analysis, writing—review and editing); AB (writing—review and editing); CR (writing—review and editing); LS (writing—review and editing); JO (writing—review and editing); GO (principal investigator, conceptualization, funding acquisition, writing—review and editing); CN (conceptualization, project administration, writing—review and editing); All authors have read and approved the manuscript, and fulfil all four criteria for authorship.

## Abstract

**Objective:** This study aims to compare how HIV-positive and HIV-negative women in a remote sub-country in Uganda access health services to inform consideration of potential HIV and HPV-based cervical cancer screening integration at the community level.

**Methods:** This cross-sectional study recruited women living in the South Busoga District Reserve from January to August 2023. Women were eligible if they were aged 30 to 49 years old, had no history of cervical cancer screening or treatment, had no previous hysterectomy, and could provide informed consent. Participants completed a survey administered by village health teams, which included questions on HIV status, demographics, healthcare access, and services received. The data was analyzed using bivariate descriptive statistics, including chi-square and Fisher’s exact tests.

**Results:** Among the 1437 participants included in the analysis, 8.8% were HIV-positive. The majority of the respondents were between 30-39 years of age, were married, had received primary education or higher, and were farmers. The majority of women in both groups had accessed outreach visits (HIV-positive = 89.0%, HIV-negative = 85.8%) and health centres (HIV-positive = 96.1%, HIV-negative = 80.2%). The most commonly received services among both groups of women at outreach visits and health centres were immunization and antenatal care, respectively.

**Conclusion:** Our study demonstrated that there were no significant differences in healthcare access between HIV-positive and HIV-negative women in rural Uganda. Additionally, the high usage of healthcare services by women living with HIV suggests that the integration of cervical cancer and HIV screening may facilitate early detection and prevention of cervical cancer among this population. This can reduce the burden of disease in Uganda and further contribute to the World Health Organization’s initiative to eradicate cervical cancer.

## Introduction

Cervical cancer is the most common cancer among women in low- and middle-income countries (LMICs). (1, 2). Despite cervical cancer being almost entirely preventable and treatable (2), LMICs face a significant burden of disease as prevention efforts such as vaccination and screening services are often limited (3, 4). Exacerbating the disproportionate burden of cervical cancer in LMICs, women are often diagnosed at advanced stages and are unable to receive effective treatment due to limited-service accessibility and availability (5). Uganda has one of the highest incidence rates of cervical cancer in the world, with 56.2 reported cases per 100,000 women (3).

In 2020, the World Health Organization (WHO) announced its global strategy to eliminate cervical cancer (6), with a key target of 70% of women screened with high-performance screening tests by the age of 35 (6). Although cervical cancer is one of the leading causes of morbidity and mortality among women in Uganda, less than 10% of women have been screened (5), highlighting the urgent need for an accessible screening program across the country. Many women in LMICs also experience personal, logistical, and healthcare system barriers, leading to a low rate of screening uptake (8). Self-collected human papillomavirus (HPV)-based cervix screening has been proposed as an effective and cost-efficient alternative strategy to overcome existing barriers to cervical cancer screening and further reduce cervical cancer rates in LMICs (9,10).

HPV-based cervical cancer screening allows for the identification of high-risk HPV, which is a primary cause of cervical cancer (1). Self-collection offers a less invasive, low-barrier approach to cervical screening that can be performed by women on their own, in the privacy of their homes or wherever convenient (11). It has been shown to be as accurate as cervical samples collected by clinicians and more sensitive than visual inspection with acetic acid (VIA), which is currently the most commonly used method to screen women in LMICs (11-13). With these benefits, HPV-based screening may allow for increased access and coverage of high-quality cervical cancer screening in LMICs, such as Uganda (12).

Women living with human immunodeficiency virus (HIV) are six times more likely to develop invasive cervical cancer compared to women without HIV infections, making cervical cancer prevention among this population a key priority (1,14,15). Additionally, women living with HIV face many barriers to accessing health services such as financial burdens (16), limited awareness (17) availability of specialized care (16), and social stigma (16,18). Ensuring access to cervical cancer screening for HIV-positive women may contribute to reducing HIV-attributable cervical cancer cases, and further alleviate the heightened burden of disease in Uganda (19). To effectively facilitate cervical cancer prevention amongst this population, the integration of HIV and HPV-based cervical cancer screening may be beneficial. Sarah Maria et al. found that less than half of HIV-positive women in urban Uganda had ever been screened for cervical cancer (20), a number that is likely to be lower in rural areas. Integration could ultimately reduce barriers, specifically among women living in rural Uganda who experience additional challenges such as long distances to health facilities and lack of health information access (21). Additionally, literature from LMICs demonstrates that integration can improve the utilization of both HIV care and cervical cancer screening services, alleviate stigma, and enhance the overall quality of care received by patients (22).

The potential for successful and effective integration of HIV and HPV-based cervical cancer screening services in Uganda relies on multiple factors, including understanding how women with HIV currently access health services compared to women without HIV. However, there is limited research on this topic, particularly in rural areas. The objective of this study is to compare how HIV-positive and HIV-negative women in a remote sub-country in Uganda access health services to inform consideration of potential HIV and HPV-based cervical cancer screening integration at the community level.

## Methods

### Design, Setting, and Study Population

This cross-sectional study is part of a pragmatic cluster-randomized trial in Malongo, a rural and remote sub-county within the Mayuge district of Uganda focused on HPV-based self-collection for cervical cancer screening. The survey data used for this study is part of a baseline survey given to participants at enrollment in the pragmatic trial to assess knowledge and attitudes about cervical cancer and screening. Women were recruited door-to-door by village health teams (VHTs) between January 23 to August 24, 2023 from eleven villages in the South Busoga Forest Reserve in Malongo. Women were eligible for the study if they were aged 30-49 years old, had not previously been screened for cervical cancer, did not have a previous hysterectomy, and could provide informed consent. VHTs administered the survey on REDCap software (23) using electronic tablets to ensure secure data collection. All questions were read aloud in Lusoga or English, based on the language preference of the participant.

### Survey

The survey tool used in this study is informed by the Improving Data for Decision Making in Global Cervical Cancer Programs Toolkit-Part 2 (IDCCP) (24). The survey included questions on participant demographics, medical history, knowledge about cervical cancer, and perceived barriers to screening.

The explanatory variable used in the study was self-reported HIV status. Participants of the survey were asked if they had ever tested positive for HIV and responded with either yes, no, or prefer not to say. Women who answered yes (HIV-positive) or no (HIV-negative) were included in the study.

Several variables were used to assess the outcome of access to health services. First, we asked women whether they had attended either a healthcare outreach visit or visited a health centre. Healthcare outreach visits included organized community health events or other instances where health services were available at the community level. Health centres refer to established clinics or hospitals that deliver healthcare services to patients. Response options for both variables were yes, no, and do not know. If a woman indicated do not know, it was recoded as no. Women were asked what services they received at the outreach visit and health centre visit, if applicable. Response options for both variables were antenatal, HIV, family planning, immunization, and other. Additionally, if applicable, women were asked about how they were recruited to a health outreach visit. Response options included: VHTs, radio, and other, which combined community leader, community member, family/friend, and other. Women were also asked, if applicable, how they traveled to health centers. Response options included: walking, boda boda, and other.

Secondly, several variables related to cervical cancer screening were included to assess the potential integration of HIV care and cervical cancer screening. These variables included: prior awareness of cervical cancer, the barriers to cervical cancer screening (did not know how/where to get the test, embarrassment, too expensive, didn’t have time, clinic too far away, poor service quality, afraid of the procedure, afraid of social stigma, cultural beliefs, family member would not allow, other, do not know), and perceived importance of early cervical cancer detection.

The following demographic variables were included in the analysis: survey language, age, education level, occupation, relationship status, partner education level, age at first pregnancy, and number of pregnancies.

### Data analysis

Bivariate descriptive statistics were used to analyze the findings of the survey data collected in the study. All missing values were included in the results. Chi-square and Fisher’s exact test were used to determine statistically significant variables (25). P-values ≤ 0.05 were considered statistically significant. R Studio (R 4.3.0) was used to conduct the analysis (26).

### Ethics Statements

Ethics approval was obtained from the University of British Columbia Children’s and Women’s Research Ethics Board (H22-01634), the Uganda Cancer Institute Research Ethics Committee (UCI-2022-56), and the Uganda National Council for Science and Technology. All participants provided informed consent for their inclusion in the study.

## Results

Overall, 1437 participants completed the survey, of which 127 women were HIV-positive (8.8%). The majority of the respondents in both groups (HIV-negative and HIV-positive) were between 30-39 years of age, had obtained primary education or more (HIV-positive n=76, 59.8%; HIV-negative n=900, 68.7%; p=0.123), were married (HIV-positive n=107, 84.3%; HIV-negative n=1136, 86.7%; p=0.865), and were farmers (HIV-positive n=99, 78.0%; HIV-negative n=1117, 85.3%; p=0.06). Additional information on the demographic characteristics of the survey respondents is provided in **Table 1**.

**Table 1.** Demographics stratified by HIV status.

|  | Overall<br>N = 1437<br>*n (%)* | HIV-positive<br>N = 127<br>*n (%)* | HIV-negative<br>N = 1310<br>*n (%)* | p-value |
| --- | --- | --- | --- | --- |
| **Survey Language** |  |  |  | < 0.001 |
| English | 38 (2.6) | 14 (11.0) | 24 (1.8) |  |
| Lusoga | 1399 (97.4) | 113 (89.0) | 1286 (98.2) |  |
| **Age** |  |  |  | 0.143 |
| 30-34 | 612 (42.6) | 47 (37.0) | 565 (43.1) |  |
| 35-39 | 319 (22.2) | 35 (27.6) | 284 (21.7) |  |
| 40-44 | 249 (17.3) | 17 (13.4) | 232 (17.7) |  |
| 45-49 | 257 (17.9) | 28 (22.0) | 229 (17.5) |  |
| **Education** |  |  |  | 0.123 |
| No Education | 451 (31.4) | 50 (39.4) | 401 (30.6) |  |
| Primary Education or more | 976 (67.9) | 76 (59.8) | 900 (68.7) |  |
| Missing | 10 (0.7) | 1 (0.8) | 9 (0.7) |  |
| **Occupation** |  |  |  | 0.060 |
| Farmer | 1216 (84.6) | 99 (78.0) | 1117 (85.3) |  |
| Other | 178 (12.4) | 21 (16.5) | 157 (12.0) |  |
| Missing | 43 (3.0) | 7 (5.5) | 36 (2.7) |  |
| **Relationship Status** |  |  |  | 0.865 |
| Married/In a relationship | 1243 (86.5) | 107 (84.3) | 1136 (86.7) |  |
| Single | 116 (8.1) | 12 (9.4) | 104 (7.9) |  |
| Separated/Divorced/Widowed | 77 (5.4) | 8 (6.3) | 69 (5.3) |  |
| Missing | 1 (0.1) | 0 (0.0) | 1 (0.1) |  |
| **Partner Education** |  |  |  | 0.813 |
| No Education | 189 (13.2) | 17 (13.4) | 172 (13.1) |  |
| Primary Education or more | 1037 (72.2) | 89 (70.1) | 948 (72.4) |  |
| Missing | 211 (14.7) | 21 (16.5) | 190 (14.5) |  |
| **Age at first pregnancy** |  |  |  | 0.16 |
| < 16 | 262 (43.3) | 28 (35.4) | 234 (44.5) |  |
| ≥ 16 | 313 (51.7) | 45 (57.0) | 268 (51.0) |  |
| Missing | 25 (4.1) | 4 (5.1) | 21 (4.0) |  |
| **Number of pregnancies** |  |  |  | 0.875 |
| 1-6 | 40 (2.8) | 28 (35.4) | 255 (48.5) |  |
| 7-12 | 256 (17.8) | 21 (16.5) | 235 (17.9) |  |
| ≥ 13 | 1141 (79.4) | 103 (81.1) | 1038 (79.2) |  |

When participants were asked about their history of attendance at healthcare outreach visits, the majority of women, regardless of their HIV status, responded yes to attending (HIV-positive n=113, 89.0%; HIV-negative n=1124, 85.8%; p=0.596). Among the 1237 women who attended a health outreach visit, immunization was reported as the most commonly accessed service for both groups (HIV-positive n=59, 46.5%; HIV-negative n=551, 42.1%; p=0.388), followed by antenatal care (HIV-positive n=47, 37.0%; HIV-negative n=517, 39.5%; p=0.655). When asked about how they were informed of health care outreach visits, the vast majority in both groups indicated VHT (HIV-positive n=102, 80.3%; HIV-negative n=1051, 80.2%; p=0.036).

More women attended a health centre than healthcare outreach visits (HIV-positive n=122, 96.1%; HIV-negative n=1267, 96.7%; p=0.289). Among the 1389 women who attended a health center, antenatal care was reported as the most commonly accessed service for both groups (HIV-positive n=79, 62.2%; HIV-negative n=839, 64.0%; p=0.752) followed by HIV services for HIV-positive women (HIV-positive n=41, 32.3%; p<0.001) and family planning for HIV-negative women (HIV-negative n=311, 23.7%; p=0.749) (**Table 2**). The respondents in both groups reported relying mainly on walking as their primary means of transportation to reach health centres (HIV-positive n=84, 66.1%; HIV-negative n=839, 64.0%; p=0.373).

**Table 2.**
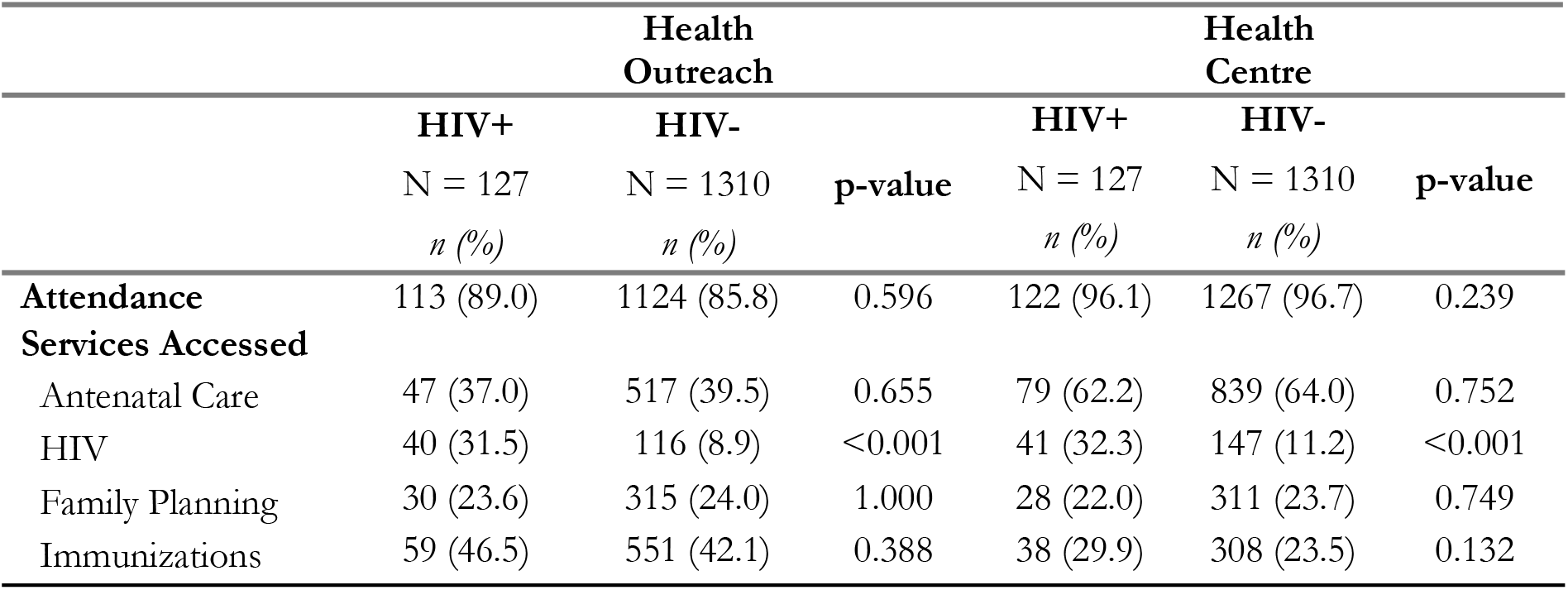
Results of survey on attendance to healthcare services stratified by HIV status.

The majority of women across both groups answered that they had prior awareness of cervical cancer (HIV-positive n= 103, 81.1%; HIV-negative n= 1137, 86.8%; p=0.321). Furthermore, the majority believed that early detection was important (HIV-positive n= 116, 91.3%; HIV-negative n= 1185, 90.6%; p=0.01). When asked about the barriers to screening (**Figure 1**), the most common response among both groups was not knowing where to get tested (HIV-positive n= 37, 29.1%; HIV-negative n= 522, 39.8%) followed by the cost (HIV-positive n= 36, 28.3%; HIV-negative n= 356, 27.2%), and prohibitive distance to the clinic (HIV-positive n= 19, 15.0%; HIV-negative n= 191, 14.6%) (p=0.023).

**Figure 1.**
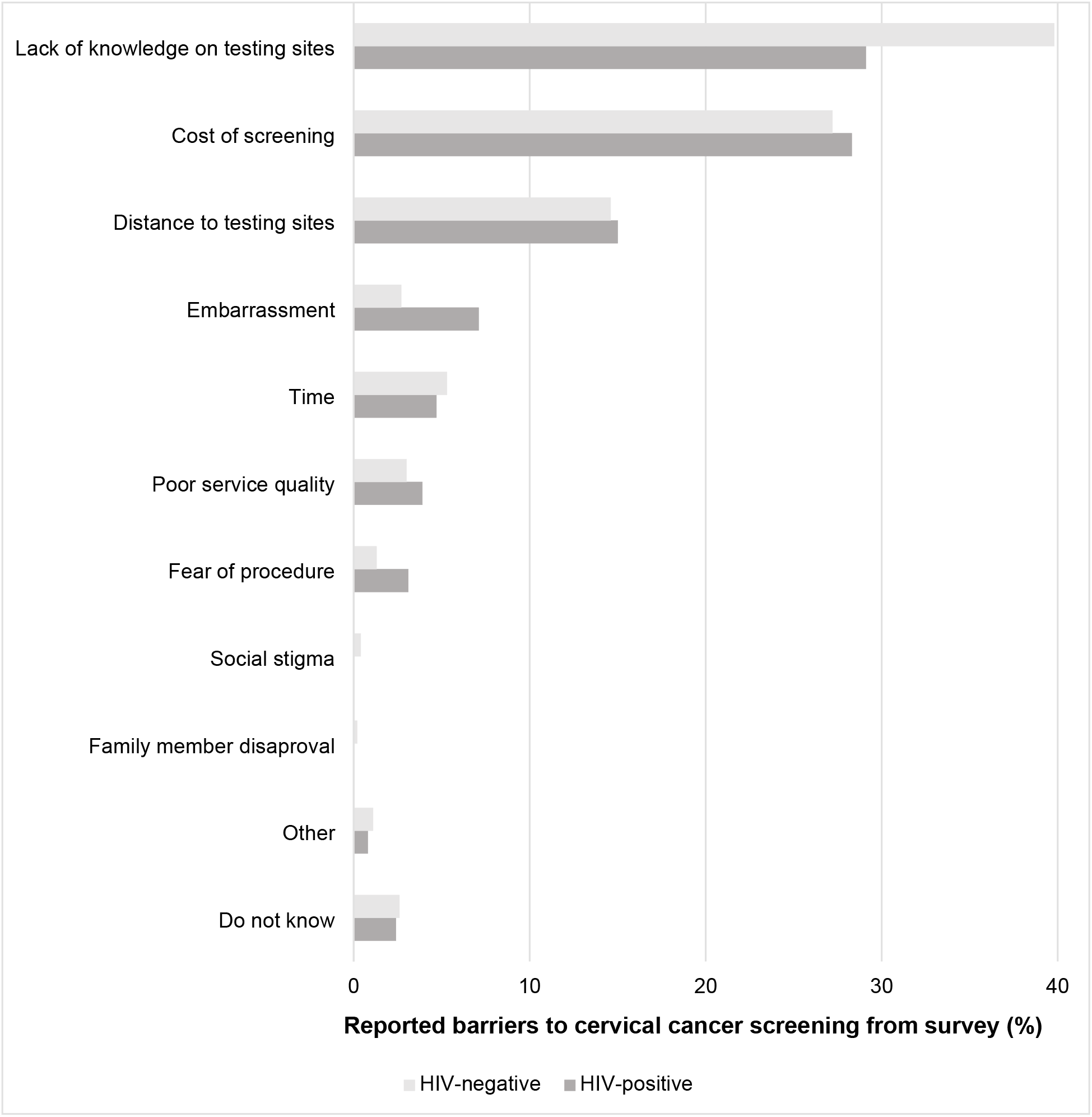
Common barriers to cervical cancer screening stratified by HIV status

## Discussion

This study aimed to assess the differences in healthcare access by HIV status. We found that the majority of women in both HIV-positive and HIV-negative groups attended health care outreach visits and health centres and utilized similar services at both.

The majority of HIV-positive and HIV-negative women had prior awareness of cervical cancer and understood the importance of early cervical cancer detection. The lack of cervical cancer knowledge is likely not a barrier to accessing screening among never-screened women in Malongo. Research conducted in Zimbabwe (27), Ghana (28), and Western Uganda (29) on cervical cancer knowledge and attitudes of women found that women generally had good levels of understanding of HPV, cervical cancer, and screening (27-29). However, findings demonstrated that knowledge and information on cervical cancer did not lead to behaviour change for screening, as represented by the low rates of screening among the women in these countries (27, 29). Additionally, there was no significant difference in knowledge between HIV-positive and HIV-negative women (28). In Uganda, cervical cancer screening rates are less than 10% (5), despite the findings of our study indicating widespread awareness of cervical cancer in women, regardless of HIV status. There is a need for programming to bridge the gap between knowledge and behaviour change to ensure that awareness is translated into increased screening to alleviate the burden of disease among women in Uganda (30).

The majority of women in both groups who attended health outreach visits were informed about these visits by VHTs. Current literature demonstrates the critical role that VHTs play in disseminating health information and facilitating access to health services in Uganda (31). As a primary point of contact for health and social services, VHTs assist community members in understanding available healthcare services and options and mobilize individuals in the community to actively participate in accessible health programs (31, 32). Their role in the healthcare of Uganda can help overcome barriers to participation and encourage community members to access services available to them. The findings from our study along with the evidence provided in the literature suggest that VHTs could play a critical role in raising awareness of HPV-based cervical cancer screening and available screening options in both HIV-positive and negative populations. Moreover, VHTs may contribute to the promotion and establishment of the successful integration of HIV and HPV-based cervical cancer screening services in rural communities.

Our study indicated that all women had a history of pregnancy and that a high proportion of HIV-positive and HIV-negative women had received antenatal services. Studies that have shown a high rate of attendance to antenatal care among pregnant women in Uganda (34). In addition to the notable overlap of the age of women attending antenatal care and the age of women recommended to be screened for cervical cancer (35), pregnancy is a period where women experience heightened medical supervision and prioritize their own health conditions (36). Furthermore, women attending antenatal care show a strong commitment to adhere to follow-up visits and care instructions throughout their pregnancy (37). Therefore, the antenatal period may provide an opportunity for increased uptake of HPV-based cervical cancer screening, both in HIV-negative and HIV-positive women in Uganda (36).

Given that HIV-positive women are at a higher risk for developing cervical cancer, it is critical to prioritize and cervical cancer prevention amongst this population (1,13,14). Investigating the differences in healthcare access and service utilization between HIV-positive and HIV-negative women allows for a better understanding of additional barriers that women living with HIV may be facing in accessing cervical cancer prevention services. This study highlighted that there were no significant differences in health service access and usage between HIV-positive and HIV-negative women. This demonstrates the ways in which women living with HIV are generally similarly engaged with the healthcare system in rural Uganda compared to HIV-negative women. This may further suggest that health inequities faced by women living with HIV are not being exacerbated by barriers to accessing and utilizing available health services, suggesting that lack of access does not fully explain the low uptake rates of cervical cancer screening reported among HIV-positive women (20). These findings can inform additional investigation into the barriers that prevent the uptake of cervical cancer screening in this population., as well as the development of a comprehensive HPV-based cervical cancer screening in Uganda that effectively targets women regardless of their HIV status. Further, this may lead to increased access to reproductive services among both HIV-positive and negative women in rural and remote communities, allowing for expanded health access for all. Integrated, concurrent screening for HIV and HPV may be more effective in the early detection and treatment of pre-cancerous lesions among women living with HIV, reducing the burden of disease that these women face (40).

The findings of this study are strengthened by the experience and the expertise of the study team. Additionally, study recruitment and survey administration were also conducted door-to-door, meaning women were involved in the study from the safety and comfort of their own homes. Despite the strengths of this study, some limitations exist. As the prevalence of HIV was much higher in the women who were surveyed in comparison to the population data of women living in Uganda, the results of this study may not be generalizable (41). This difference in HIV prevalence may be due to self-reporting of HIV status, which could impact the results of the study. Similarly, the collected survey data was based on self-reported information, which may be subject to response biases that influence the results of the study (42). As women in the study responded to the survey questions to VHTs verbally, this may have resulted in social desirability bias (43).

The results from our study demonstrate that women in remote Uganda had high attendance to health services offered at both outreach visits and health centres, with no significant differences between HIV-positive and HIV-negative women. Further research is needed on how to effectively integrate HIV and HPV-based cervical cancer screening programs into rural health systems at the community level.

## Data Availability

Data is not available for access as it contains human data that contains potentially identifying information about participants.

## References

1. Cervical cancer [Internet]. World Health Organization; 2020 [cited 2023 Jun 20]. Available from: https://www.who.int/news-room/fact-sheets/detail/cervical-cancer

2. Mitchell S, Ogilvie G, Steinberg M, Sekikubo M, Biryabarema C, Money D. Assessing women’s willingness to collect their own cervical samples for HPV testing as part of the ASPIRE cervical cancer screening project in Uganda. International Journal of Gynecology & Obstetrics. 2011;114(2):111–5.

3. Gottschlich A, Payne BA, Trawin J, Albert A, Jeronimo J, Mitchell-Foster S, et al. Community-integrated self-collected HPV-based cervix screening in a low-resource rural setting: a pragmatic, cluster-randomized trial. Nat Med. 2023 Apr;29(4):927–35.

4. Mitchell SM, Sekikubo M, Biryabarema C, Byamugisha JJK, Steinberg M, Jeronimo J, et al. Factors associated with high-risk HPV positivity in a low-resource setting in sub-Saharan Africa. American Journal of Obstetrics and Gynecology. 2014 Jan;210(1):81.e1-81.e7.

5. Hull R, Mbele M, Makhafola T, Hicks C, Wang S, Reis R, et al. Cervical cancer in low and middle-income countries (Review). Oncol Lett. 2020 Jun 19;20(3):2058–74.

6. WHO Global strategy to accelerate the elimination of cervical cancer as a public health problem [Internet]. World Health Organization; 2020 [cited 2023 Jun 29]. Available from: https://www.who.int/publications/i/item/9789240014107

7. Mungo C, Barker E, Randa M, Ambaka J, Osongo CO. Integration of cervical cancer screening into HIV/AIDS care in low-income countries: a moral imperative. Ecancermedicalscience. 2021 May 20;15:1237.

8. Teng FF, Mitchell SM, Sekikubo M, Biryabarema C, Byamugisha JK, Steinberg M, et al. Understanding the role of embarrassment in gynaecological screening: a qualitative study from the ASPIRE cervical cancer screening project in Uganda. BMJ Open. 2014 Apr 1;4(4):e004783.

9. Oketch SY, Kwena Z, Choi Y, Adewumi K, Moghadassi M, Bukusi EA, et al. Perspectives of women participating in a cervical cancer screening campaign with community-based HPV self-sampling in rural western Kenya: a qualitative study. BMC Women’s Health. 2019 Jun 13;19(1):75.

10. Megersa BS, Bussmann H, Bärnighausen T, Muche AA, Alemu K, Deckert A. Community cervical cancer screening: Barriers to successful home-based HPV self-sampling in Dabat district, North Gondar, Ethiopia. A qualitative study. PLOS ONE. 2020 Dec 11;15(12):e0243036.

11. Bakiewicz A, Rasch V, Mwaiselage J, Linde DS. “The best thing is that you are doing it for yourself” – perspectives on acceptability and feasibility of HPV self-sampling among cervical cancer screening clients in Tanzania: a qualitative pilot study. BMC Women’s Health. 2020 Dec;20(1):65.

12. Brandt T, Wubneh SB, Handebo S, Debalkie G, Ayanaw Y, Alemu K, et al. Genital self-sampling for HPV-based cervical cancer screening: a qualitative study of preferences and barriers in rural Ethiopia. BMC Public Health. 2019 Jul 31;19(1):1026.

13. Woo YL, Gravitt P, Khor SK, Ng CW, Saville M. Accelerating action on cervical screening in lower- and middle-income countries (LMICs) post COVID-19 era. Prev Med. 2021 Mar;144:106294.

14. Mitchell SM, Pedersen HN, Eng Stime E, Sekikubo M, Moses E, Mwesigwa D, et al. Self-collection based HPV testing for cervical cancer screening among women living with HIV in Uganda: a descriptive analysis of knowledge, intentions to screen and factors associated with HPV positivity. BMC Women’s Health. 2017 Dec;17(1):4.

15. Mungo C, Barker E, Randa M, Ambaka J, Osongo CO. Integration of cervical cancer screening into HIV/AIDS care in low-income countries: a moral imperative. Ecancermedicalscience. 2021 May 20;15:1237.

16. Bouabida K, Chaves BG, Anane E. Challenges and barriers to HIV care engagement and care cascade: viewpoint. Front Reprod Health. 2023 Jul 20;5:1201087.

17. Luseno WK, Wechsberg WM, Kline TL, Ellerson RM. Health Services Utilization Among South African Women Living with HIV and Reporting Sexual and Substance-Use Risk Behaviors. AIDS Patient Care STDS. 2010 Apr;24(4):257–64.

18. Lofgren SM, Tsui S, Atuyambe L, Ankunda L, Komuhendo R, Wamala N, et al. Barriers to HIV Care in Uganda and Implications for Universal Test-and-Treat: A Qualitative Study. AIDS Care. 2022 May;34(5):597–605.

19. Stelzle D, Tanaka LF, Lee KK, Ibrahim Khalil A, Baussano I, Shah ASV, et al. Estimates of the global burden of cervical cancer associated with HIV. The Lancet Global Health. 2021 Feb;9(2):e161–9.

20. Sarah Maria N, Olwit C, Kaggwa MM, Nabirye RC, Ngabirano TD. Cervical cancer screening among HIV-positive women in urban Uganda: a cross sectional study. BMC Women’s Health. 2022 May 10;22(1):148.

21. Ndejjo R, Mukama T, Musabyimana A, Musoke D. Uptake of Cervical Cancer Screening and Associated Factors among Women in Rural Uganda: A Cross Sectional Study. PLoS One. 2016;11(2):e0149696.

22. Huchko MJ, Maloba M, Nakalembe M, Cohen CR. The time has come to make cervical cancer prevention an essential part of comprehensive sexual and reproductive health services for HIV-positive women in low-income countries. Journal of the International AIDS Society. 2015;18(6S5):20282.

23. Patridge EF, Bardyn TP. Research Electronic Data Capture (REDCap). J Med Libr Assoc [Internet]. 2018 Jan 12 [cited 2023 Jun 23];106(1). Available from: http://jmla.pitt.edu/ojs/jmla/article/view/319

24. World Health Organization. Improving data for decision-making: a toolkit for cervical cancer prevention and control programmes [Internet]. Geneva: World Health Organization; 2018 [cited 2023 Jun 23]. 289 p. Available from: https://apps.who.int/iris/handle/10665/279420

25. Andrade C. The P Value and Statistical Significance: Misunderstandings, Explanations, Challenges, and Alternatives. Indian J Psychol Med. 2019;41(3):210–5.

26. R Core Team. R: A Language and Environment for Statistical Computing [Internet]. Vienna, Austria; 2023. Available from: https://www.R-project.org/

27. Fitzpatrick M, Pathipati MP, McCarty K, Rosenthal A, Katzenstein D, Chirenje ZM, et al. Knowledge, attitudes, and practices of cervical Cancer screening among HIV-positive and HIV-negative women participating in human papillomavirus screening in rural Zimbabwe. BMC Women’s Health. 2020 Jul 25;20(1):153.

28. Stuart A, Obiri-Yeboah D, Adu-Sarkodie Y, Hayfron-Benjamin A, Akorsu AD, Mayaud P. Knowledge and experience of a cohort of HIV-positive and HIV-negative Ghanaian women after undergoing human papillomavirus and cervical cancer screening. BMC Women’s Health. 2019 Oct 23;19(1):123.

29. Usman IM, Chama N, Aigbogun Jr EO, Kabanyoro A, Kasozi KI, Usman CO, et al. Knowledge, Attitude, and Practice Toward Cervical Cancer Screening Among Female University Students in Ishaka Western Uganda. Int J Womens Health. 2023 Apr 14;15:611– 20.

30. Mukama T, Ndejjo R, Musabyimana A, Halage AA, Musoke D. Women’s knowledge and attitudes towards cervical cancer prevention: a cross sectional study in Eastern Uganda. BMC Womens Health. 2017 Jan 31;17(1):9.

31. Perry S, Fair CD, Burrowes S, Holcombe SJ, Kalyesubula R. Outsiders, insiders, and intermediaries: village health teams’ negotiation of roles to provide high quality sexual, reproductive and HIV care in Nakaseke, Uganda. BMC Health Services Research. 2019 Aug 13;19(1):563.

32. Musinguzi LK, Turinawe EB, Rwemisisi JT, de Vries DH, Mafigiri DK, Muhangi D, et al. Linking communities to formal health care providers through village health teams in rural Uganda: lessons from linking social capital. Human Resources for Health. 2017 Jan 11;15(1):4.

33. Turinawe EB, Rwemisisi JT, Musinguzi LK, de Groot M, Muhangi D, de Vries DH, et al. Selection and performance of village health teams (VHTs) in Uganda: lessons from the natural helper model of health promotion. Human Resources for Health. 2015 Sep 7;13(1):73.

34. Babughirana G, Gerards S, Mokori A, Nangosha E, Kremers S, Gubbels J. Maternal and newborn healthcare practices: assessment of the uptake of lifesaving services in Hoima District, Uganda. BMC Pregnancy and Childbirth. 2020 Nov 11;20(1):686.

35. Filade TE, Dareng EO, Olawande T, Fagbohun TA, Adebayo AO, Adebamowo CA. Attitude to Human Papillomavirus Deoxyribonucleic Acid-Based Cervical Cancer Screening in Antenatal Care in Nigeria: A Qualitative Study. Front Public Health. 2017;5:226.

36. Kuczborska K, Kacperczyk-Bartnik J, Wolska M, Pluta M, Bartnik P, Dobrowolska-Redo A, et al. Secondary cervical cancer prevention in routine prenatal care — coverage, results and lessons for the future. Ginekologia Polska. 2019;90(7):396–402.

37. Brun-Micaleff E, Coffy A, Rey V, Didelot MN, Combecal J, Doutre S, et al. Cervical cancer screening by cytology and human papillomavirus testing during pregnancy in french women with poor adhesion to regular cervical screening. Journal of Medical Virology. 2014;86(3):536–45.

38. Tchounga B, Boni SP, Koffi JJ, Horo AG, Tanon A, Messou E, et al. Cervical cancer screening uptake and correlates among HIV-infected women: a cross-sectional survey in Côte d’Ivoire, West Africa. BMJ Open. 2019 Aug;9(8):e029882.

39. White HL, Meglioli A, Chowdhury R, Nuccio O. Integrating cervical cancer screening and preventive treatment with family planning and HIV-related services. Int J Gynecol Obstet. 2017 Jul;138:41–6.

40. Ninsiima M, Nyabigambo A, Kagaayi J. Acceptability of integration of cervical cancer screening into routine HIV care, associated factors and perceptions among HIV-infected women: a mixed methods study at Mbarara Regional Referral Hospital, Uganda. BMC Health Serv Res. 2023 Apr 3;23(1):333.

41. Uganda Population-Based HIV Impact Assessment [Internet]. PHIA Project; 2022 [cited 2023 July 2]. Available from: https://phia.icap.columbia.edu/wp-content/uploads/2022/08/UPHIA-Summary-Sheet-2020.pdf

42. Rosenman R, Tennekoon V, Hill LG. Measuring bias in self-reported data. Int J Behav Healthc Res. 2011 Oct;2(4):320–32.

43. Althubaiti A. Information bias in health research: definition, pitfalls, and adjustment methods. JMDH. 2016 May;211.

